# Adopting an American framework to optimize nursing admission documentation in an Australian health organization

**DOI:** 10.1101/2021.06.15.21258998

**Authors:** Danielle Ritz Shala, Aaron Jones, Greg Fairbrother, Melissa Baysari

**Author notes:** Corresponding author: Danielle Ritz Shala, BSN, RN, MSc Health Data Science, MACN, Sydney Local Health District Nursing and Midwifery Services/Health Informatics, Level 4, Building 89, 50 Missenden Road, Camperdown NSW 2050. Declarations of interest: none.

## Abstract

**Objective:** Apply and modify the American Essential Clinical Dataset (ECD) approach to optimize the data elements of an electronic nursing admission assessment form in a metropolitan Australian local health district.

**Materials and Methods:** We used the American ECD approach but made modifications. Our approach included 1) a review of data, 2) a review of current admission practice via consultations with nurses, 3) a review of evidence and policies, 4) workshops with nursing and informatics teams in partnership with the eMR vendor, and 5) team debrief sessions to consolidate findings and decide what data elements should be kept, moved, or removed from the admission form.

**Results:** Of 165 data elements in the form, 32% (n=53) had 0% usage, while 25% (n=43) had 100% usage. Nurses’ perceptions of the form’s purpose varied. Eight policy documents specifically prescribed data to be noted at admission. Workshops revealed risks of moving or removing data elements, but also uncovered ways of streamlining the form. Consolidation of findings from all phases resulted in a recommendation to reduce 91% of data elements.

**Discussion:** Application of a modified ECD approach allowed the team to identify opportunities for significantly reducing and reorganizing data elements in the eMR to enhance the utility, quality, visibility, and value of nursing admission data.

**Conclusion:** We found the modified ECD approach effective for identifying data elements and work processes that were unnecessary and duplicated. Our findings and methodology can inform improvements in nursing clinical practice, information management, and governance in a digital health age.

## INTRODUCTION

The use of electronic medical records (eMRs) for inpatient care has steadily increased in Australia, with governments continuing to invest in their widespread adoption.[1-3] In New South Wales (NSW), Australia’s most populous state, a statewide electronic medical record project was implemented in 2013 with a focus on streamlining clinical documentation for nursing and midwifery, including completion of mandatory assessments and observations during the admission process.[4] The acute care admission process is a significant event in the patient care episode. It requires specific documentation for planning and execution of nursing care and a clear overview of the patient for all members of the multidisciplinary team.[5,6]

It has been acknowledged that over-documentation in eMRs does not add value to care team processes.[6] In a US study that examined data utilization for medical decision-making during admission, it was found that 25% of the clinical data available in the eMR was never used, and only 33% was used more than 50% of the time by admitting physicians.[7] eMR systems indisputably enable the capture of and access to large amounts of patient data, but optimizing data item content choice and presenting this in a way that best supports clinicians’ decision-making processes, is much less clearly understood and agreed upon.[5]

While efforts to improve eMR utilization have proliferated in recent years, a systematic review showed that the use of electronic health records increased nursing documentation time by 22-44%.[8] Recent Swedish findings also indicated that eMRs failed to reflect nursing practice with nurses altering their routines to fit the system, rather than the system being tailored to suit nursing practice.[5] Such findings highlight a need to improve eMR systems so that they better support nursing clinical workflows and optimize documentation times.

In 2018, an American collaborative developed and applied a systematic approach to simplify documentation during the acute care inpatient admission process and identify a minimal required dataset - the Essential Clinical Dataset (ECD) approach.[6] The ECD approach improves nursing care and data quality by: (1) allowing nurses to focus on pertinent/essential information, (2) reducing unnecessary documentation, and (3) reinforcing/augmenting nursing time for direct patient care. Application of the ECD approach in the USA significantly reduced documentation item content by 49% in a collaborative of 12 healthcare organizations [6] and reduced documentation time for nurses by 72% in a separate 600-bed academic medical center.[9] In this current study, we set out to apply the ECD approach to optimize nursing admission documentation in a metropolitan Australian local health district. In particular, we aimed to apply a modified ECD approach to identify data elements that could be moved or removed from the nursing adult admission assessment (AAA) form in the eMR.

## MATERIALS AND METHODS

### Setting

This study was conducted in a Sydney-based health district which encompasses three large tertiary referral hospitals (>2,000 bed total capacity), an inpatient rehabilitation facility and a network of community health services. From April to June 2020, the district had a total of 35,144 admitted patient episodes, and 81,606 acute overnight bed days.[10] Electronic patient information in the hospitals is primarily managed via the Cerner Millennium eMR.

### The Essential Clinical Dataset Approach

The American ECD approach included a literature review, US-federal regulatory review, and environmental scans of the eMR to determine utilization rates of data elements completed by nurses. Our approach was guided by the logic and spirit of the American ECD approach, but we were required to make a number of changes because not all metrics used in the American approach were available in NSW, and we faced time and resource constraints posed by the COVID-19 pandemic. Importantly, we also avoided a top-down approach and instead involved end-users of the eMR system and AAA form, and eMR and nursing informatics teams in informal consultations and workshops. Five phases were completed in sequence as illustrated in Table 1. A comparison between our approach and the American ECD approach is outlined in Appendix A.

**Table 1:** Our interpretation of the ECD approach

|  |  |
| --- | --- |
| Phase 1<br>**Review of Data** | Review of AAA form compliance and data element utilization using eMR data |
| Phase 2<br>**Review of Practice** | Review of current admission practice and workflow via consultation with nurses |
| Phase 3<br>**Review of Policy** | Review of evidence and policy relevant to nursing admission and documentation |
| Phase 4<br>**ECD Workshop** | Workshop with nursing and informatics teams to rationalize data elements to be retained and understand impacts and user perceptions of moving or removing data elements |
| Phase 5<br>**Post Workshop Debrief** | Consolidation of findings from all phases to arrive at recommendations for essential admission data elements to maintain in the eMR |

#### Phase 1: Review of AAA form compliance

A retrospective review of all AAA forms (n=92,957) completed between November 2018 to November 2019 in the shared eMR domain of two local health districts was performed to examine form completion among nurses. In particular, the usage rate of each data element (n=165 data elements) in each of the 19 sections of the AAA form was determined, as were the mandatory and conditional data elements in the AAA form. Utilization was defined as the actual number of times the data element was documented by a nurse.[6] Data was extracted by Cerner from the shared eMR domain. Descriptive statistics were used to analyze the utilization of each data element in the AAA form. The timeliness of AAA form completion was determined in a separate analysis of adult inpatient admissions (n=37,512) and calculated as the interval between date and time of admission and form completion.[11]

#### Phase 2: Review of practice

Consultations were conducted with 11 Clinical Nurse Educators from adult inpatient units and Emergency Departments across three hospitals in November and December 2019. The discussions focused on nursing admission workflow, information gathered during admission and associated documentation in the eMR. Educators were encouraged to share their experiences and provide examples. Handwritten notes were taken during discussions and these were then reviewed by two team members to identify key points.

#### Phase 3: Review of policy

A policy review was undertaken to determine policy and evidence directly related to questions in the AAA form. To provide a clear scope of review and ensure direct relevance of policies to the data elements in question, data were extracted from policies only if they clearly stated what information should be collected upon a patient’s admission. Clinical Nurse Consultants and Department Directors acted as subject matter experts (SMEs) for this aspect of the project by providing specific policies directly related to the AAA data elements. A total of 13 key policy documents were reviewed.

#### Phase 4: ECD Workshop

Findings from the data, policy, and practice review phases were presented and discussed with nursing staff in ECD Workshops to rationalize data elements to be kept, moved, or removed from the AAA form. This also provided an avenue for the ECD team to explore staff insights, suggestions, concerns, and potential risks of form changes or data element recommendations. Four workshop sessions lasting one hour each were held online via Zoom. An online questionnaire was emailed to participants after the first workshop which asked participants to indicate which data elements should be kept, moved, or removed from the AAA form. Results were tallied prior to the second workshop and used to prioritize data elements for discussion in the workshop sessions.

Workshops were facilitated by the local district, in partnership with Cerner. Screenshots of the powerform, along with the corresponding utilization rates of the data elements were presented. The facilitators emphasized the International ECD Collaborative practice recommendations for reviewing data elements, which includes challenging traditional practice, reminding staff to “think outside the box,” explaining why data needs to be collected, and identifying other appropriate eMR sections.[6] Scenarios and examples were provided to demonstrate concepts where relevant. Data collected during workshops included suggestions for streamlining the form, as well as the risks and benefits associated with removing or moving items from the AAA.

#### Phase 5: ECD Team debrief and summative review

Team debrief sessions were held after each workshop to discuss areas of improvement, data element recommendations, and to identify priorities for the succeeding workshops. At the end of Workshop 4, the ECD team, comprising the district’s Chief Nursing and Midwifery Information Officer, ECD Coordinator, Applications Specialist, and Cerner Australia’s Chief Nursing Officer, Senior Nursing Clinical Consultants and Solution Architect, reviewed the data elements and recommendations discussed in the workshops. Items that required further investigation, clarification, or were not discussed at workshops due to time constraints were also discussed, and a final proposal was made regarding which data elements to keep, move or remove from the AAA form.

## RESULTS

### Phase 1: Review of AAA form compliance

Fifty three data elements (32%) had nil usage relative to the form, while 43 data elements (25%) had 100% usage. The remaining 69 data elements (42%) had variable usage rates. Of the 42 data elements with perfect usage rates, 32 were mandatory fields, which are required to be documented before the AAA form can be signed off. The non-mandatory data elements with perfect usage were either calculations or data automatically completed by the eMR system based on answers to other data elements. Of the 53 data elements which were never used, 18 were part of the patient belongings section, 15 part of a mental status exam/cognitive assessment (Abbreviated Mental Test Score) section, and 20 were part of the risk screening and assessment section of the AAA form (see table 2).

**Figure 1:**
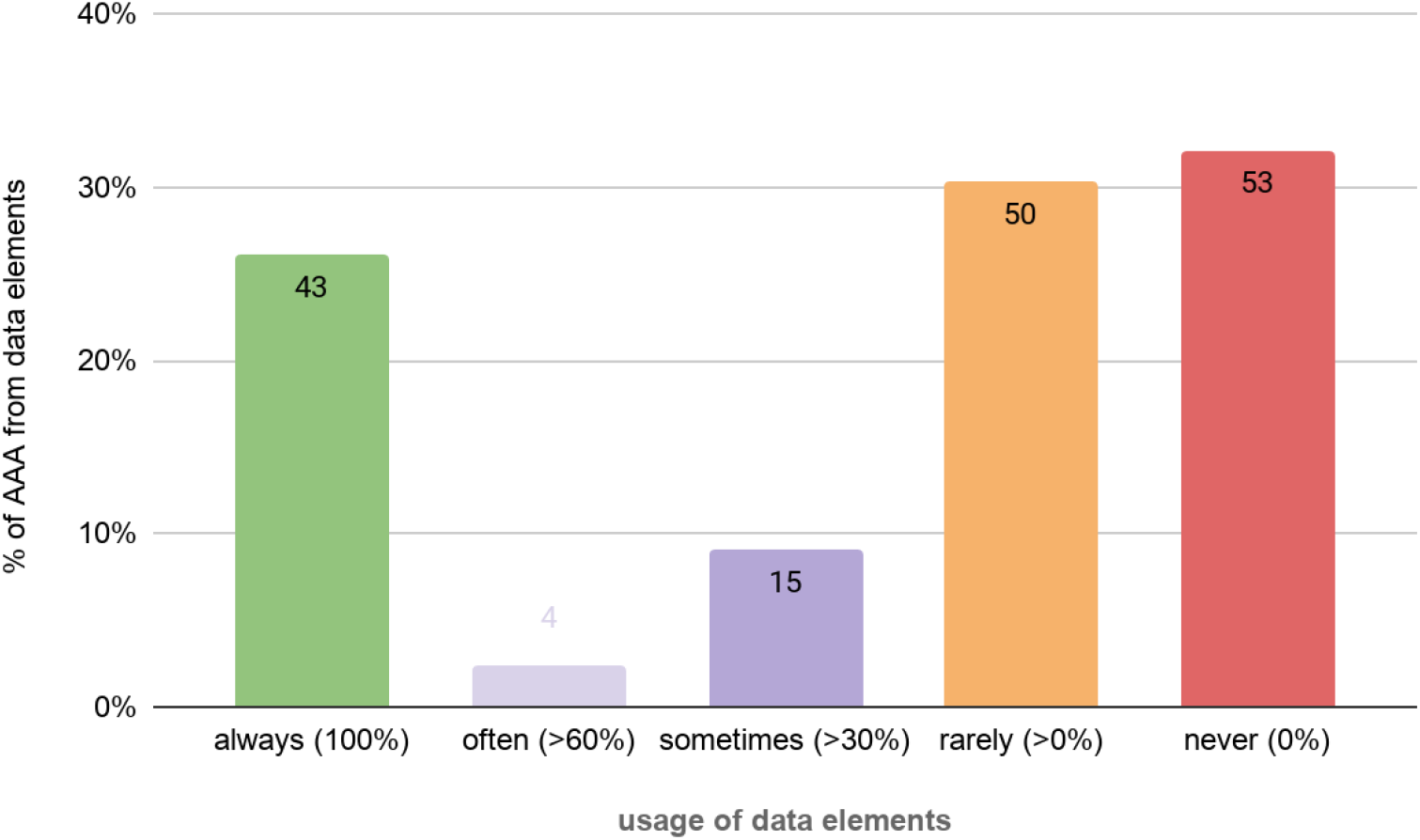
Usage of 165 AAA form data elements.

**Table 2:** Usage rates of data elements in each AAA section

|  |  | *Completion rates of data elements* |  |  |  |  |  |
| --- | --- | --- | --- | --- | --- | --- | --- |
|  | *AAA form section* | **Always**<br>**(100%)** | **Often**<br>**(≥60%)** | **Sometimes**<br>**(≥30%)** | **Rarely**<br>**(≥30%)** | **Never**<br>**(0%)** | **Total data**<br>**elements** |
| 1 | Admission Risk Assessment Screen | 1 | 1 | 10 | 3 | 1 | **16** |
| 2 | Allergies and Adverse Reactions | - | - | - | - | 1 | **1** |
| 3 | Alerts | - | - | - | 1 | 1 | **2** |
| 4 | Screening for Risk | 13 | 3 | 4 | 7 | 8 | **35** |
| 5 | Mobility and Transfer | 6 | - | - | - | 1 | **7** |
| 6 | Nutrition and Toileting | 6 | - | - | 1 | 1 | **8** |
| 7 | Mental Status and Medications | 6 | - | - | 6 | 3 | **15** |
| 8 | Abbreviated Mental Health Test | - | - | - | - | 13 | **13** |
| 9 | Risk Scores | 7 |  | 1 | 1 | 4 | **13** |
| 10 | Waterlow Prevention Equipment V2 | - | - | - | 4 |  | **4** |
| 11 | Discharge Risk Assessment | 4 | - | - | 4 | 1 | **9** |
| 12 | Patient Belongings & Valuables Checklist | - | - | - | 13 | 2 | **15** |
| 13 | Patient Belongings, Glasses | - | - | - | 1 | 2 | **3** |
| 14 | Patient Belongings, Dentures | - | - | - | 2 | 3 | **5** |
| 15 | Patient Belongings, Hearing Aids | - | - | - | 1 | 2 | **3** |
| 16 | Patient Belongings, Contact Lenses | - | - | - | - | 3 | **3** |
| 17 | Patient Belongings, Mobile/Elec Dev | - | - | - | 1 | 1 | **2** |
| 18 | Patient Valuables, Money | - | - | - | 3 | 3 | **6** |
| 19 | Patient Valuables, Jewellery | - | - | - | 2 | 3 | **5** |
|  | **Total data elements** | **43** | **4** | **15** | **50** | **53** | **165** |

In a separate analysis of inpatient encounters, 78% (n=22,953) of AAA forms were completed within the first 24 hours of admission, 13.3% (n=3,910) between 24-72 hours from admission, and 8.7% (n=2558) beyond 72 hours from admission.[11]

### Phase 2: Review of practice

The key points from consultations with nursing staff were grouped into: 1) The role of documentation in the nursing admission process, (2) Barriers to electronic documentation of admission, and (3) Perceived advantages and disadvantages of the AAA form

#### Role of documentation in the nursing admission process

Each nurse consulted outlined the key steps of an admission process for their unit. While there were slight variations in each account and the sequence of steps, key elements were identified, namely: handover of the patient from ED or theatre (where the admission was not booked), patient identification and orientation to unit, taking vital signs/observations, checking of IV fluids and lines, checking of medications, nursing assessments, creating a nursing admission progress note, informing multidisciplinary teams and making referrals if necessary. These discrete steps were reported to happen simultaneously/dynamically and involve documentation in different sections of the eMR. Nurses mentioned that admission information is found and documented across different places, including the electronic AAA form, progress notes, nursing handover form, and printed handover sheets.

Nurses expressed different views about the purpose of the AAA form. Some nurses saw the purpose beyond care planning, and suggested that it is also used for auditing and incident management. Others viewed the form as an electronic checklist which can guide and/or remind staff about steps and tasks to perform as part of the admission process. For example, a nurse explained that the “Patient orientation to ward” section, which is a checkbox list enumerating tasks expected from nurses upon admission (e.g. orienting patients with call bell, toilets, providing hospital brochures, etc.) serves as a useful prompt for nurses, while others thought that a checklist was not necessary for these items, as they are considered part of routine nursing admission care.

In addition to this, completing the AAA form was often described as a “tick-box exercise” by nurses. Some expressed concern that the form is perceived as a task that needs to be complied with, but once completed, the admission task is done. A nurse stated that for most staff, completing the AAA is “something that they have to do more than something they value.” Many nurses expressed that when filling out eMR forms such as the AAA, most staff tend to complete only the mandatory fields.

#### Barriers to electronic documentation of admission

Nurses described the following key barriers to electronic documentation of admission: (1) timing of admission, (2) unit acuity and staffing, and (3) nursing experience. Some nurses indicated that it was not always an appropriate time to have a discussion with a patient. Completing the AAA form was reported to be challenging (and so sometimes not done) if patients are admitted towards the commencement of the night shift, as nurses preferred patients to rest at this time-point. Asking multiple or repetitive questions of patients in the middle of the night, or at the time of arrival to the unit may be inappropriate, particularly if patients are unwell. Nurses also provided examples relating to times of peak, focused nursing activity, such as medication administration, handover, or the arrival of further new admissions. Nurses explained that these times are not optimal for performing a risk assessment, or completing comprehensive documentation, the result being that nursing admission documentation would often be delayed.

Nurses identified unit acuity and staffing as factors that could also influence nursing admission documentation. They reported that if the unit is busy, they may not have enough time to complete all documentation relating to admission. Similarly, if there are not enough staff on the floor, other tasks relating to direct patient care and procedures may be prioritized over admission documentation. The level of nursing experience was also mentioned as a factor associated with the time it takes to complete a nursing admission and relevant documentation. Clinical Nurse Educators indicated that on average, it takes approximately 15-20 minutes for a senior nurse to complete a nursing admission with documentation, while junior nursing staff could take up to 40 minutes.

#### Perceived advantages and disadvantages of the AAA form

The main perceived advantage of the current AAA form was its convenience, as it pulls together other forms required for admission (e.g. pressure injury and falls risk forms) into a single electronic form. One nurse described the AAA form as a “one-stop shop,” making it easier for nursing staff to complete admission documentation. Despite this, disadvantages were also noted as staff described the questions in the form as “too scattered” and “repetitive.” The form was also perceived to be inflexible, as it does not allow a user to access other sections of the eMR while it is open/once started. Repetition of information and duplication of tasks were frequently cited as problems.

### Phase 3: Review of policy

The policy review highlighted the Australian National Standards for Quality in Health Safety (NSQHS) Communicating for Safety and Comprehensive Care standards as the principal regulatory basis for ECD. We also identified eight key policies that specifically prescribed data to be collected during admission (see Appendix B). The policies recommended information about allergies, medications, height and weight, patient belongings, and a transfer of care risk assessment be documented at admission or at point of presentation to hospital.

### Phase 4: ECD Workshop

When asked to indicate which data elements should be kept, moved, or removed from the AAA form in the pre-workshop survey, most of the participants opted to keep most of the data elements in the form. During workshop discussions, a key risk of moving or removing data elements from the AAA form was identified to be the flow on effect this would have to other sections of the eMR. For example, when pregnancy-related data elements were discussed, a participant explained that removing some of these elements may affect the clinical ranges in the observations chart for a pregnant patient who comes to hospital with a non-maternity presentation (e.g. fracture, trauma, etc.). Another risk identified by participants was the potential decrease in compliance among staff if data elements are moved or removed. Nurses were concerned that the completion of admission-related tasks such as the pressure injury and falls risk forms may decrease if the data elements are not contained in a single form.

To reduce the number of questions and streamline the form, workshop participants suggested removing closely related or repetitive data elements and compressing them into a single question. Leveraging the form’s conditional logic functionality would mean that questions are shown only if relevant to a previous response. For example, the patient belongings section, which comprises more than 20 questions in the AAA could be replaced with a single yes/no question, “does the patient come with any belongings?” The patient belongings form would then only be triggered if the answer is yes. Participants felt that this would significantly reduce the data elements and sections in the AAA form, and make the form appear less busy and cluttered with questions. The same was suggested for cognition-related data elements, where multiple data elements would only be triggered if the staff answered yes to a single question.

### Phase 5: ECD Team debrief: Summative review

After reviewing data from Phases 1-4, the ECD team agreed that only 14 (9%) of the 165 data elements should be maintained in the AAA form. As shown in Table 3, it was recommended that the majority (n=93, 57%) of elements be removed from the current form either due to low utilization or the duplication of these elements in other eMR sections, and nearly a third (n=45, 27%) be moved and collected through existing sections of the eMR.

**Table 3.** Recommendations for maintenance, movement or removal of data elements in AAA form

| AAA FORM SECTIONS |  | RECOMMENDATIONS FOR DATA ELEMENTS |  |  |  |  |  |  |  |  |  |
| --- | --- | --- | --- | --- | --- | --- | --- | --- | --- | --- | --- |
|  |  | Keep |  | Move |  | Remove |  | NA |  | TOTAL |  |
| Section | Section Name | n | % | n | % | n | % | NA | % | n | % |
| 1 | Admission Risk Assessment Screen | 3 | 19 | 1 | 6 | 11 | 69 | 1 | 6 | 16 | 100 |
| 2 | Allergies and Adverse Reactions | 1 | 100 | - | - | - | - | - | - | 1 | 100 |
| 3 | Alerts | 1 | 50 | - | - | - | - | 1 | 50 | 2 | 100 |
| 4 | Screening for Risk | 2 | 6 | 14 | 40 | 15 | 43 | 4 | 11 | 35 | 100 |
| 5 | Mobility and Transfer | - | - | 6 | 86 | - | - | 1 | 14 | 7 | 100 |
| 6 | Nutrition and Toileting | - | - | 6 | 86 | - | - | 1 | 14 | 7 | 100 |
| 7 | Mental Status and Medications | 2 | 13 | 2 | 13 | 10 | 67 | 1 | 7 | 15 | 100 |
| 8 | Abbreviated Mental Health Test | - | - | - | - | 13 | 100 | - | - | 13 | 100 |
| 9 | Risk Scores | - | - | 12 | 92 | - | - | 1 | 8 | 13 | 100 |
| 10 | Waterlow Prevention Equipment V2 | - | - | 4 | 100 | - | - | - | - | 4 | 100 |
| 11 | Discharge Risk Assessment | 4 | 44 | - | - | 4 | 44 | 1 | 11 | 9 | 100 |
| 12 | Patient Belongings & Valuables Checklist | 1 | 7 | - | - | 13 | 87 | 1 | 7 | 15 | 100 |
| 13 | Patient Belongings, Glasses | - | - | - | - | 3 | 100 | - | - | 3 | 100 |
| 14 | Patient Belongings, Dentures | - | - | - | - | 5 | 100 | - | - | 5 | 100 |
| 15 | Patient Belongings, Hearing Aids | - | - | - | - | 3 | 100 | - | - | 3 | 100 |
| 16 | Patient Belongings, Contact Lenses | - | - | - | - | 3 | 100 | - | - | 3 | 100 |
| 17 | Patient Belongings, Mobile/Elec Dev | - | - | - | - | 2 | 100 | - | - | 2 | 100 |
| 18 | Patient Valuables, Money | - | - | - | - | 6 | 100 | - | - | 6 | 100 |
| 19 | Patient Valuables, Jewellery | - | - | - | - | 5 | 100 | - | - | 5 | 100 |
| **TOTAL** |  | **14** | **9** | **45** | **27** | **93** | **57** | **12** | **7** | **164** | **100** |

## DISCUSSION

The ECD method revealed that the current AAA form includes a large number of unused data elements. Of the 165 data elements, only a quarter are always completed or used, while almost two thirds never or rarely used. More importantly, application of our modified ECD approach allowed the team to identify optimization opportunities, specifically 93 data elements that could be removed and 45 data elements that could be moved from the current form. This would reduce the total data elements in the form by 92%, allowing nurses to focus on essential clinical information relating to a patient’s admission. As most of the AAA data elements were found to be present in other sections of the eMR, the proposed optimization would minimize duplication in data entry, clarify the appropriate eMR sections where information should be recorded, and increase the visibility of nursing admission data for nursing and multidisciplinary care teams.

Although the current study focused on Australian nursing admission data and processes, the findings are comparable to those from the North American approach. For example, our rates of data element utilization are similar to those reported in a US medical admission study where only 33% of clinical data available in the eMR was used more than 50% of the time by physicians for decision making at admission.[7] Our recommended reduction in data elements from the AA form also mirrors results from the North American ECD with one organization arriving at a reduction of 91% of data elements, and an average reduction in data elements for the entire cohort of 49%.[6]

We found the ECD approach very effective for identifying work processes and elements that were unnecessary and duplicated. Many of these processes were likely to be present when paper-based systems were in place. Transitioning from paper to electronic documentation is typically initially focused on digitizing medical records and supporting paper-based work practices to continue electronically. Deviations or innovations are kept to a minimum in the early stages of implementation because practitioners are unfamiliar with the new technology and are learning about its capabilities for the first time.[12] However, once clinicians and nurses have been well inducted in the practice of electronic documentation, there is more opportunity to review current electronic documentation practices and policy, and explore more innovative and efficient approaches to support clinical care. Emphasis should be placed on supporting nurses’ decision making but electronic systems should go beyond checklists and merely transferring documentation from paper to the computer,[13] to simplify, streamline, or automate tasks.[14]

Redundant data elements were identified in the existing adult admission assessment form with nurses questioning why some data elements are repeated in multiple eMR locations. This result is consistent with the findings of a study on Australian aged care admission forms, where items referring to the same concept had different names in different form formats.[15] The same study showed that multiple items designed to capture information had overlapping meanings and nurses found these vague, leading to low completion rates. Data relating to similar concepts scattered in the eMR can also lead to increased cognitive load, misrepresentations, and uncertainty about the source of truth for patient care planning.[7,12,16] Clinicians could be spending time unnecessarily identifying the most relevant data for clinical decision making, potentially selecting the wrong data, and missing important information, thereby increasing patient safety risks.[12]

Nurses held various views about the purpose of the AAA form and these multiple functions could have contributed to the high number of data elements included in the initial design of the form, and the low completion rates. While nurses agreed that the form guides patient care planning, other purposes such as checklist reminders, audits, and incident management were also identified. This is not surprising as nursing documentation comprises technical, scientific, legal, and ethical aspects that provide health systems with records to audit nursing actions and estimate the quality of patient care.[13] However, this same variation in purpose could have contributed to the addition of data elements that extend beyond the main objective of care planning, making the form longer. Without careful oversight, this can be problematic, as an increased number of data elements across different nursing forms including admission has been associated with poor comprehensiveness, increased documentation workload, confusion, and incomplete documentation among nurses.[15,16]

We found very limited guidance in policy documents on particular items to be collected during an admission, suggesting that the data elements in the AAA could have originated from a combination of historical and socio-technical processes. From a historical point of view, studies have described processes of transitioning from paper to electronic documentation in nursing and healthcare in general.[13,17,18] Implementing electronic documentation in nursing has standardized various formats of paper-based admission forms in Australia, some of which were directly automated into an electronic format.[15] From a socio-technical point of view, form content and design are often informed by working groups with physicians, nurses, clinical managers or administrators, health informaticians, and technical staff as key players.[6,13,15,19] In a study which reviewed admission data elements to compare the quality of aged care admission forms in Australia, it was acknowledged that different formats of admission forms were developed and validated by experienced nursing managers in each organization, and the nursing knowledge captured across all items was assumed to be valuable and respected. Similarly, an American case study describing the transition of a medical center from paper to electronic documentation [18] outlined how clinical medical leaders designed electronic templates corresponding to paper note headings for admission and progress notes because the templates provided by the eMR vendor were deemed inadequate for the context of their clinical setting. To ensure the local context and workflows were understood and incorporated into our ECD process, we followed the American ECD model by including representatives from the eMR vendor and the district’s nursing informatics and eMR teams, but also engaged with clinical nurse educators during consultations and included these end-users as workshop participants. These clinical leaders were a key driver in deciding the content and structure of eMR forms, ensuring the AAA form “fit” their clinical practice. Their involvement, particularly in the workshop discussions, allowed the team to understand nuances that were not evident with the objective data derived from the eMR environmental scan and utilization rates.

### Limitations

In-person workshops were not possible and the time for nursing staff engagement was strictly limited due to the COVID-19 pandemic. We recognize that this was a stressful period across the health system and participants’ opportunity to focus on workshop materials, reflect on ECD principles, and engage in online workshops could have been limited. The team mitigated this limitation through the pre-workshop online data elements survey and encouraging staff to provide feedback post-workshop via email, MS teams, or phone call. Not all AAA data elements were discussed with nurses during the four workshop sessions, and recommendations for the outstanding items were finalized by the ECD team.

## CONCLUSION

Adopting the ECD methodology in Australia has uncovered opportunities for improving nursing clinical practice, information management, and governance. Reducing and streamlining data elements in the adult admission assessment form is recommended to enhance the quality, utility, visibility, and value of nursing data. The project has established the value of modifying an international framework, analyzing local district data, and collaborating with eMR, clinical, academic, and eMR vendor teams in reviewing data, practice, and policy for eMR optimization. End-user involvement is crucial in bridging the gap between technology and clinical practice and in ensuring that proposed changes are aligned with clinical workflows and best practice. The localized approach can be used by other health organizations for optimizing nursing documentation, and can potentially be shared with other medical and clinical disciplines for future optimization work. This could transform nursing and clinical data into meaningful information, and maximize technology to support communication, efficiency, and timeliness for patient care.

## Data Availability

N/A

## ACKNOWLEDGMENT

We would like to acknowledge the support of the executives, contribution of managers, and participation of staff from the local health district’s Nursing and Midwifery Services, Health Informatics Teams, and ICT Services in this innovation initiative. We would also like to thank Cerner USA and Cerner Australia for their support in this project.

