## Supplementary file_APPENDIX A for "Adopting an American framework to optimize nursing admission documentation in an Australian health organization"

Comparison of Cerner International ECD and the modified Australian ECD approaches

|  | **International ECD** | **Australian ECD** |
| --- | --- | --- |
| **PHASE 1**  ***Review of Data*** |  |  |
| Baseline metrics   - Cerner data element extract - eMR form timers   *(time elapsed from form activation to form completion)*   - eMR Click counters *(number of clicks while completing the form)* - eMR explorer Adult Admission Assessment Powerform audit report | Yes  Yes  Yes  No | Yes  Not available  Not available  Yes |
| Quantitative Analysis   - Data element utilization rates - Time from activating form to completing form - Number of clicks during form completion - Time elapsed from admission time to form completion time | Yes  Yes  Yes  No | Yes  Not available  Not available  Yes |
| **Phase 2**  ***Review of Practice*** |  |  |
| Approach | eMR environmental scans | Consultations with nursing staff (Clinical Nurse Educators and managers) in the district about admission and AAA form |
| **Phase 3**  ***Review of Policy*** |  |  |
| Resources reviewed | USA federal regulatory standards  Literature review | Australian National Standards  NSW Ministry of Health guidelines/policies  Consultation with district subject matter experts (Clinical Nurse Consultants) to provide national, state, or local district policies that specifically relate to data elements in the AAA form at point of admission  Relevant Australian and NSW health literature |
| **Phase 4**  ***ECD Workshop*** |  |  |
| ECD Collaborative workshop participants | Cerner USA  Chief Nursing Informatics Officers | Cerner Australia  Chief Nursing and Midwifery Information Officer  ECD Coordinator  Health Informatics teams  Clinical Nurse Educators  eMR applications specialist |
| Duration | 8 hours | 4 sessions, 1-hour each  (total = 4 hours) |
| Mode of delivery | Face to face workshop | Online Zoom workshops due to COVID pandemic |
| Additional preparations | Undetermined  Not included in IECD method | Workshop materials sent to participants 3 weeks before workshop (spreadsheet with AAA data element utilization rates; policy review summary)  Online ECD data elements survey completed by participants before workshop 2 |
| Quantitative data presented at workshop   - Data element utilization rates - ECD data elements online survey | Yes  Not applicable | Yes  Yes |
| Focus of workshop discussions for data elements | All data elements | Prioritized by results of ECD data elements online survey |
| Visual presentations during workshop | Screenshots of Powerform displayed on-screen during discussions  Use of tick, cross, arrow, question mark for workshop recommendations (keep, remove, move, undetermined) | Screenshots of Powerform with data element utilization rates displayed on-screen throughout discussions  Use of tick, cross, arrow, question mark for workshop recommendations (keep, remove, move, undetermined) |
| **Phase 5**  ***Post Workshop debriefs*** |  |  |
| Focus | Not applicable | ECD team consolidation of recommendations, including data elements not discussed during workshop due to time constraints |
