## Supplementary file_APPENDIX B for "Adopting an American framework to optimize nursing admission documentation in an Australian health organization"

Table of relevant policies for the AAA form

| **Policies with specific relevance to AAA data elements** (satisfied policy review criteria) | | | | |
| --- | --- | --- | --- | --- |
| **Document** | **Clinical topic** | **Year last reviewed** | **Policy Level** | **AAA section with relevant data elements** |
| NSQHS Standard 6: Communicating for Safety | Healthcare documentation | 2017 | national | all |
| NSQHS Standard 5: Comprehensive Care | Integrated Care | 2017 | national | all |
| NSQHS Standard 4: Medication Safety Standard | Medication safety | 2017 | national | 1-Admission Risk Assessment Screen; 4-Screening for Risk |
| NSQHS Standard 5: Patient Identification and Procedure Matching | Patient Identification | 2012 | national | 1-Admission Risk Assessment Screen |
| NSW Health Protecting People and Property Manual | People and Property | 2018 | state | 12-19 Patient Belongings & Valuables |
| NSW Health PD2017_041: Nutrition Care | Nutrition | 2017 | state | 4-Screening for Risk, 6-Nutrition and Toileting |
| NSW Health PD2013_043: Medication Handling in NSW Public Health Facilities | Medications | 2013 | state | 1-Admission Risk Assessment Screen; 4-Screening for Risk |
| NSW Health PD2011_015: Care Coordination: Planning from Admission to Transfer of Care in NSW Public Hospitals | Transfer of care | 2011 | state | 11-Discharge Risk Assessment |
| **Other policies** (related but did not satisfy policy review criteria) | | | | |
| **Document** | **Clinical Topic** | **Year last reviewed** | **Policy Level** | **AAA section with relevant data elements** |
| NSW Health GL2008_001 Nursing & Midwifery Clinical Guidelines - Identifying & Responding to Drug & Alcohol Issues | Drug and Alcohol | 2008 | state | 4-Screening for Risk |
| Safe and high-quality care for patients with cognitive impairment (dementia and delirium) in hospital | Dementia | 2014 | national | 4-Screening for Risk, 7-Mental Status and Medications, 8-Abbreviated Mental Health Test |
| SLHD Delirium Policy – SLHD_PCP2019_023 | Delirium | 2019 | district | 4-Screening for Risk; 7-Mental Status and Medications, 8-Abbreviated Mental Health Test |
| SLHD Pressure Injury Prevention and Management Policy SLHD_PD2017_034 | Skin Integrity | 2017 | district | 4-Screening for Risk, 6-Nutrition and Toileting, 10-Waterlow Prevention Equipment V2 |
